# High content image analysis in routine diagnostic histopathology predicts outcomes in HPV-associated oropharyngeal squamous cell carcinomas

**DOI:** 10.1101/2022.06.24.22276368

**Authors:** Jonas Hue, Zaneta Valinciute, Selvam Thavaraj, Lorenzo Veschini

## Abstract

**Objective:** Routine haematoxylin and eosin (H&E) photomicrographs from human papillomavirus-associated oropharyngeal squamous cell carcinomas (HPV+OpSCC) contain a wealth of prognostic information. In this study, we set out to develop a high content image analysis workflow to quantify features of H&E images from HPV+OpSCC patients to identify prognostic features which can be used for prediction of patient outcomes.

**Methods:** We have developed a dedicated image analysis workflow using open-source software, for single-cell segmentation and classification. This workflow was applied to a set of 567 images from diagnostic H&E slides in a retrospective cohort of HPV+OpSCC patients with favourable (n = 29) and unfavourable (n = 29) outcomes. Using our method, we have identified 31 quantitative prognostic features which were quantified in each sample and used to train a neural network model to predict patient outcomes. The model was validated by k-fold cross-validation using 10 folds and a test set.

**Results:** Univariate and multivariate statistical analyses revealed significant differences between the two patient outcome groups in 31 and 16 variables respectively (*P*<0.05). The neural network model had an overall accuracy of 78.8% and 77.7% in recognising favourable and unfavourable prognosis patients when applied to the test set and k-fold cross-validation respectively.

**Conclusion:** Our open-source H&E analysis workflow and model can predict HPV+OpSCC outcomes with promising accuracy. Our work supports the use of machine learning in digital pathology to exploit clinically relevant features in routine diagnostic pathology without additional biomarkers.

## Introduction

The worldwide incidence of HPV-associated oropharyngeal squamous cell carcinoma (HPV+OpSCC) has been steadily increasing over the past two decades and is projected to continue to rise over the next twenty years^1,2^. In parallel with the alarming incidence rise of this disease, it is now established that HPV+OpSCCs have on average better survival outcomes compared to site and stage-matched HPV negative counterparts^3^. The robust evidence of more favourable outcomes in HPV+OpSCC has led to recent calls for treatment de-escalation in this group of patients, with several clinical trials currently in progress^4^. However, it is also recognised that not all HPV+OpSCC patients will benefit from treatment de-escalation since a small but significant proportion of these tumours demonstrate disease progression or recurrence similar to HPV-independent oropharyngeal squamous cell carcinoma (HPV-OpSCC)^5-7^. Therefore, accurately predicting patients who will benefit from treatment de-escalation is key to improving their quality of life while at the same time ensuring optimal radical dose regimens in cases likely to progress or recur.

Several potential prognostic factors in HPV+OpSCC have recently been described, including tumour cytomorphology^8,9^, density^10^ and immunoprofile of tumour infiltrating lymphocytes (TILs)^11^, plasma cell^12^ and macrophage infiltrates^12^, as well as gene methylation^13^ and genomic heterogeneity^14^. Evaluating these biomarkers is a promising strategy to select patients for treatment de-escalation in clinical trials. However, evaluation of biomarkers often requires complex laboratory testing and interpretation which is costly, lack widespread availability and is overall beyond the scope of routine diagnostic histopathology services. Furthermore, the reason for unfavourable clinical outcomes is likely to be multifactorial^15^ and individual biomarkers could fail to fully capture this complexity. Accounting for multiple variables including cytomorphology, intratumoural morphologic heterogeneity, together with spatial relationships between cancer cells and the immune microenvironment are likely to improve our ability to accurately identify patients with favourable prognosis. However, dedicated tools and workflows are currently unavailable.

Within this context, we developed an automated workflow for quantitative high content image analysis (HCA) of diagnostic haematoxylin and eosin (H&E)-stained sections of HPV+OpSCC using open-source software with little computational requirements. We measured TILs, stromal plasma cells, tumour nuclear features, morphological heterogeneity of tumour cells and the spatial relationship between TILs and tumour cells. The overall aim of the current study was to assess the value of HCA to identify potentially prognostic features in routine H&E slides and to develop a neural network model based on our features to predict outcomes in HPV+OpSCC patients.

## Patients and Methods

### Patient Selection

Cases were selected from a previously characterised cohort of 231 patients HPV+OpSCC patients treated at Guy’s & St Thomas’ NHS Foundation Trust between January 2005 and December 2017^10^. HPV positivity was defined as strong and diffuse nuclear and cytoplasmic staining for p16 immunohistochemistry and diffuse or punctate signal by HPV DNA in-situ hybridisation according to current guidelines^16^. All 29 patients with unfavourable outcomes (UO), (defined as death from disease or local, regional, or distant recurrence within 5 years) were identified and included in the current study. An additional 29 patients with favourable outcomes (FO), (defined as disease-free at 5 years) were randomly selected using a random number generator. All patients were anonymised prior to image acquisition and data analysis. A further 15 patients for a workflow development group were randomly selected from the remaining cohort after excluding the 58 UO and FO patients. This group was used to develop the image analysis workflow to ensure the workflow was robust enough to handle previously unseen images. All patient data was handled in compliance with the UK Data Protection Act. The Outer North East London Research Ethics Committee of the National Research Ethics Service gave ethical approval for this work.

### Image Acquisition

Ten separate representative digital images from archival diagnostic H&E-stained slides were acquired at 100x optical magnification for each patient. Where possible, each image contained approximately 70% tumour and 30% stroma. All images were assessed by a specialist head and neck pathologist and deemed representative of the overall diagnostic slide. A total of 567 images were acquired across the 58 patients with some patients having less than ten images due to the small biopsy size.

### Image Analysis

A dedicated image analysis workflow was developed to segment and classify individual cells and cell objects in the H&E sections (Fig 1A). See Table 1 for a summary of the software used and their respective functions. To develop the image analysis workflow, 15 patients were randomly selected after excluding the 58 aforementioned UO and FO patients. Processing was automated with scripts in QuPath and macros in ImageJ (publicly available on GitHub at https://github.com/jonashue1/HPV-OpSCC-Machine-Learning).

**Figure 1.**
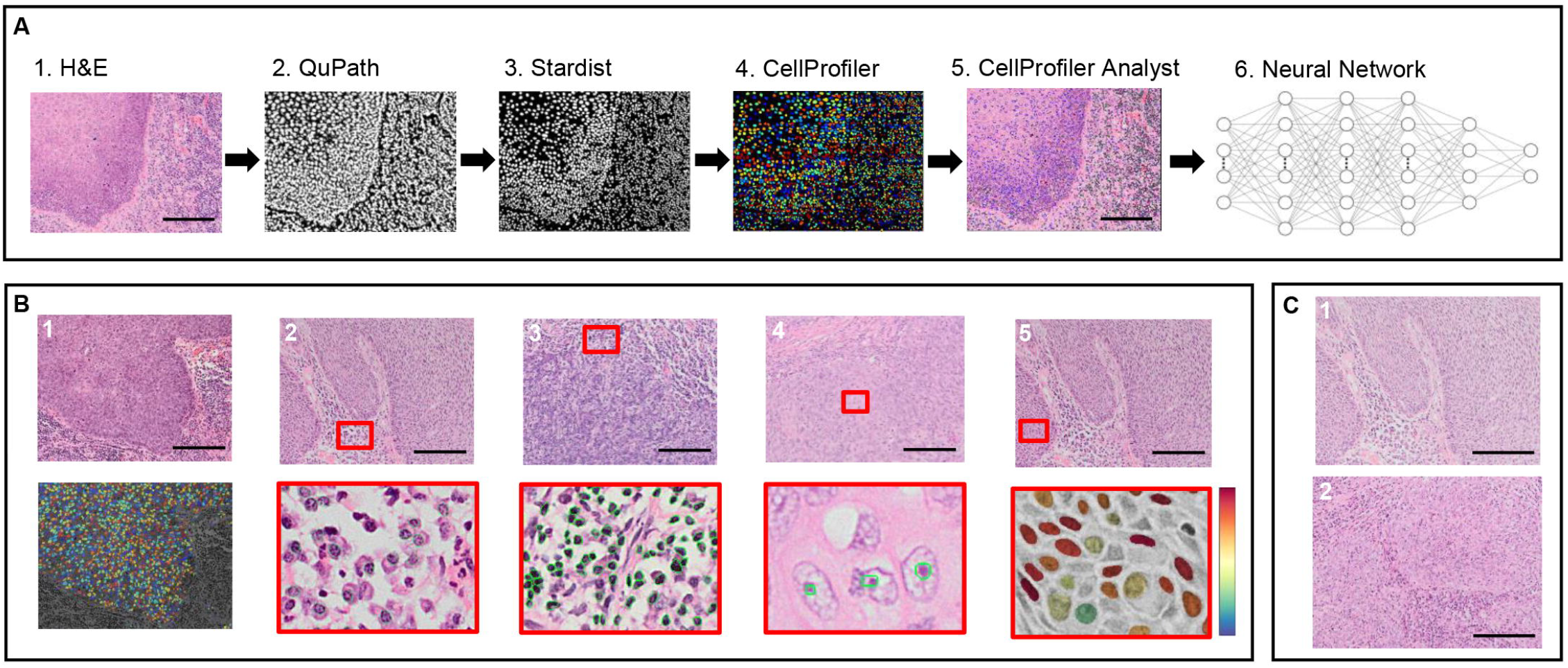
(A1-6) Overview of the image analysis workflow. H&E images (A1) analysed by ML-aided software. QuPath (A2) for initial pixel-level classification. Stardist (A3) refined nuclear segmentation for clustered objects. CellProfiler (A5). Neural network model to predict outcomes (A6). (B) Example workflow outputs of tumour nuclei (B1), plasma cells (B2), TILs (B3), nucleoli (B4) and tumour nuclear eccentricity (B5). (C) Example H&E images from favourable (C1) and unfavourable (C2) outcome groups at 100x magnification. All scale bars represent 150μm.

**Table 1.** Software used in the workflow and their respective functions.

| No. | Software | Purpose |
| --- | --- | --- |
| 1. | QuPath | Pixel-level classification with an artificial neural network pixel classifier in-built in QuPath |
| 2. | ImageJ | Unsharp mask for all images. Increasing minimum pixel intensities for faded slides. Renaming images for CellProfiler metadata. |
| 3. | Stardist | Convolutional neural network for segmentation of clustered cells |
| 4. | CellProfiler | Object-level segmentation. Feature measurements for supervised machine learning in CellProfiler Analyst. Single-cell quantification and measurements for cytomorphology spatial distribution, texture and granularity features. |
| 5. | CellProfiler Analyst | Supervised machine learning for classification of different cell types with a Fast Gentle Boosting algorithm. |
| 6. | RStudio | Univariate and multivariate data analysis. Training and validation of the prognostic neural network model. |

Image pre-processing was performed in ImageJ^17^ by applying an unsharp mask to all images. ImageJ was also used to empirically increase the minimum pixel values for faded slides from 7 patients in a supervised manner. All images were then exported to QuPath^18^ where machine learning (ML) was used to overcome the issue of stain variability of H&E images with an artificial neural network pixel-classifier. The pixel-classifier identified pixels of interest belonging to cells and cell objects for easier downstream object-based segmentation (Fig 1A2). To further improve segmentation of nuclei, Stardist was utilised as an ImageJ plugin employing a pre-trained model based on H&E images^19^ (Fig 1A3). Elaborated images were obtained and imported directly to CellProfiler^20^ for object-level segmentation (cells, tumour nuclei and nucleoli, TILs, and plasma cells) (Fig 1A4). Finally, we used an ML-based object classifier tool (CellProfiler Analyst^21^) to further discriminate cells of interest on the basis of object measurements (Fig 1A5). The various cells are classified with this model and quantified by CellProfiler. The cell classification was qualitatively reviewed by a specialist head and neck pathologist (see Fig 1B for examples of the cell classification).

An index of stromal TILs (TIL index) was calculated to attempt to account for the variation of tumour and stromal areas in each image.

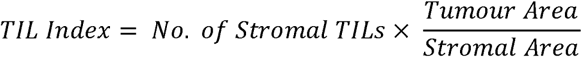

Tumour nuclear features for texture, granularity and morphology were measured in CellProfiler. The cohesiveness of the tumour was measured by the distance of each tumour cell to their first and second closest neighbours. The number of neighbours each tumour cell had was also quantified.

Intratumoural heterogeneity for nuclear morphological, textural and spatial features was quantified by the spread of the data for each respective variable and taken as the variance of the measurement per field of view. All measurements were exported to an SQLite database and imported into R.

The image analysis workflow took under 50 hours to process 567 images from 58 patients, making measurements of more than 800 000 individual tumour and stromal cells with a standard 8GB RAM computer. This averaged a processing time of approximately 51.7 minutes per patient.

### Identifying Prognostic Features

Statistical analysis was performed in R using univariate Wilcoxon’s rank sum test and multivariate logistic regression (LR) to identify variables that had a statistically significant difference between patients with unfavourable and favourable outcomes. Statistical significance was accepted at *P <* 0.05.

### Prognostic Model

The prognostic features identified were used to train a neural network to predict patient outcomes in our cohort of patients. The 567 images were split into a training and test set with 80% (n = 457) and 20% (n = 110) of images respectively. The neural network was designed with 4 hidden layers and 16, 8, 8 and 4 nodes in each respective layer. The model was further validated with a k-fold cross validation using 10 folds.

## Results

The clinicopathologic features of the patient cohort are summarised in Table 2.

**Table 2.** Clinicopathologic features of the patient cohort.

|  |  | Favourable outcome | Unfavourable outcome |
| --- | --- | --- | --- |
| Age (mean/range) |  | 62 (48-80) | 60 (47-78) |
| Sex | Male (n) | 26 | 23 |
|  | Female (n) | 4 | 5 |
| Stage | I-II (n) | 4 | 2 |
|  | III-IV (n) | 26 | 26 |

### Image Analysis Workflow

An image analysis workflow was successfully developed to segment and classify the various cell types within each biopsy. Fig 1B shows examples of the workflow being used to quantify various biopsy features.

### Statistical analysis of selected features reveals prognostic features

Quantitative measurements of the variables considered in this study are summarised in Table 3. Univariate analysis revealed that 31 variables (Fig 2) in our study were significantly different in FO versus UO patients demonstrating the value of these factors for patient stratification. TILs, regardless of tissue compartment, were more abundant in FO patients than UO patients. Plasma cell counts were also higher in the FO group with a median of 65 (97.25) versus 27 (53.5). Form factor, eccentricity, compactness cell area and perimeter are used to quantify cytomorphology. In general, FO patients had rounder and less eccentric cells, while cell area was not significantly different between the two groups. FO patients also had tumour cells packed closer to one another with shorter distances between their neighbours. FO patients had higher values for various texture and granularity measurements of tumour nuclei. Intratumoural heterogeneity was significantly greater in UO patients for most measurements of texture, granularity, cytomorphology and spatial packing.

**Table 3.** Univariate Wilcoxon’s rank sum test and multivariable logistic regression analyses for all 34 analysed variables compared between favourable and unfavourable outcome groups. Statistical significance accepted at *P*<0.05.

|  | Variable | Favourable Median | Favourable IQR | Unfavourable Median | Unfavourable IQR | Univariable Wilcoxon rank sum test *P* | Multivariable Logistic Regression *P* |
| --- | --- | --- | --- | --- | --- | --- | --- |
| Immune Cell | TIL Index | 11.31 | 22.24 | 3.16 | 10.78 | **<0.001** | **0.0074** |
|  | Intratumour TILs (n) | 24 | 38 | 9 | 22 | **<0.001** | 0.5075 |
|  | Stromal TILs (n) | 35 | 71.5 | 11 | 43 | **<0.001** | **0.0037** |
|  | Total TILs (n) | 62.5 | 114.75 | 25 | 64 | **<0.001** | 0.662 |
|  | Plasma Cells (n) | 65 | 97.25 | 27 | 53.5 | **<0.001** | **0.0082** |
| Texture & Granularity | Texture Correlation | 0.1452 | 0.077 | 0.0998 | 0.0821 | **<0.001** | 0.279 |
|  | Texture Entropy | 9.7 | 0.39 | 9.68 | 0.45 | **<0.001** | **0.0068** |
|  | Texture InfoMeas1 | 0.9957 | 0.0037 | 0.9951 | 0.0046 | **0.017** | 0.2359 |
|  | Granularity 7 | 4.76 | 3.91 | 3.89 | 3.77 | **0.002** | **0.0032** |
|  | Granularity 1 | 62.87 | 10.98 | 61.25 | 13.53 | **0.006** | 0.4221 |
| Tumour Nuclear Morphology | Form Factor | 0.841 | 0.031 | 0.809 | 0.089 | **<0.001** | 0.8471 |
|  | Compactness | 1.21 | 0.06 | 1.28 | 0.2 | **<0.001** | 0.1878 |
|  | Eccentricity | 0.718 | 0.039 | 0.733 | 0.043 | **<0.001** | **0.0018** |
|  | Perimeter (Px) | 98.37 | 12.05 | 100.65 | 10.73 | **0.0052** | **0.0096** |
|  | Cell Area (Px) | 643.02 | 190.29 | 658.77 | 189.53 | 0.0821 | 0.0631 |
| Tumour Cell Spatial Distribution | Number of Neighbours | 5.81 | 0.03 | 5.8 | 0.05 | **<0.001** | 0.2811 |
|  | 1st Neighbour Distance (Px) | 36.63 | 4.2 | 37.55 | 6.68 | **<0.001** | **<0.001** |
|  | 2nd Neighbour Distance (Px) | 47.37 | 6.91 | 49.16 | 11.05 | **<0.001** | **<0.001** |
| Other Tumour Features | Tumour Cell Count | 1449.5 | 474.75 | 1334 | 590.5 | **<0.001** | 0.0874 |
|  | Image Nucleoli | 404.5 | 341.75 | 334 | 269 | **<0.001** | 0.3394 |
|  | Nucleoli Ratio | 0.269 | 0.246 | 0.27 | 0.203 | **0.0311** | 0.4019 |
| Intra-tumour Heterogeneity | Texture Correlation | 0.449 | 0.141 | 0.399 | 0.205 | **<0.001** | **0.0232** |
|  | Texture Entropy | 0.219 | 0.139 | 0.305 | 0.256 | **<0.001** | 0.9846 |
|  | Texture InfoMeas1 | 1.03E-02 | 2.12E-02 | 0.0158 | 0.03 | **0.0011** | 0.4841 |
|  | Granularity 7 | 0.0571 | 0.0894 | 0.032 | 0.1114 | **0.0016** | **0.0067** |
|  | Granularity 1 | 0.234 | 0.18 | 0.262 | 0.278 | 0.0791 | **0.0172** |
|  | Form Factor | 0.147 | 0.202 | 0.343 | 0.432 | **<0.001** | **0.0091** |
|  | Compactness | 0.0292 | 0.0442 | 0.078 | 0.1286 | **<0.001** | 0.9956 |
|  | Eccentricity | 0.483 | 0.212 | 0.496 | 0.181 | 0.1735 | 0.1382 |
|  | Perimeter | 0.129 | 0.088 | 0.186 | 0.221 | **<0.001** | **0.001** |
|  | Cell Area | 0.187 | 0.175 | 0.207 | 0.19 | **0.0012** | **0.0089** |
|  | Number of Neighbours | 0.334 | 0.195 | 0.414 | 0.31 | **<0.001** | 0.1339 |
|  | 1st Neighbour Distance | 0.0604 | 0.055 | 0.0745 | 0.0829 | **<0.001** | **0.046** |
|  | 2nd Neighbour Distance | 0.0516 | 0.0517 | 0.0615 | 0.0613 | **0.0056** | 0.1434 |

**Figure 2.**
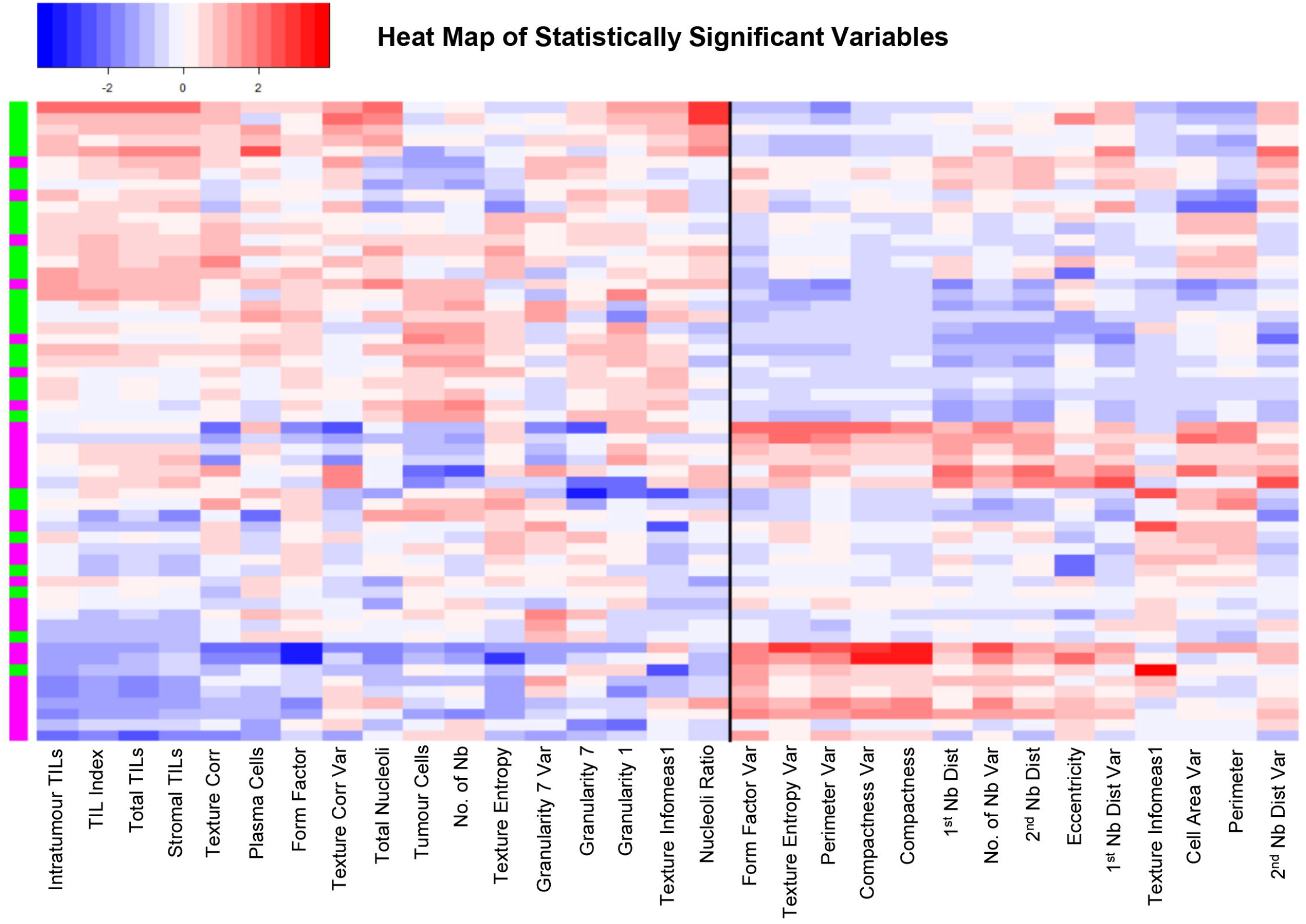
Heat map of all statistically significant variables using Euclidean distance and hierarchical clustering by complete linkage. Patients are colour-coded based on outcome, green representing favourable and magenta representing unfavourable outcomes respectively.

Overall, the heat map (Fig 2) reveals clustering of patients by outcomes. Two broad clusters appear evident from the heat map with most of FO patients clustering at the top. In general, these patients had a stronger immune infiltrate and low intratumoural heterogeneity. On the other hand, UO patients clustered at the bottom, exhibiting generally low immune infiltrate and higher intratumoural heterogeneity. Our analysis also reveals inter-tumour homogeneity for various measurements in FO patients. Conversely, patients with UO have greater intertumour heterogeneity. When these patients do not strictly follow trends, predicting their outcomes becomes increasingly difficult. Taken individually, none of the variables were predictive of the outcome as shown by the inter- and intratumoural variability. Therefore, we proceeded to analyse our data through multivariate LR and machine learning tools.

### Multivariate analysis and machine learning predicts patient outcomes

Results of the multivariate LR are summarised in Table 3. Fig 3A shows boxplots of selected variables that were statistically significant on both univariate and multivariate analyses.

**Figure 3.**
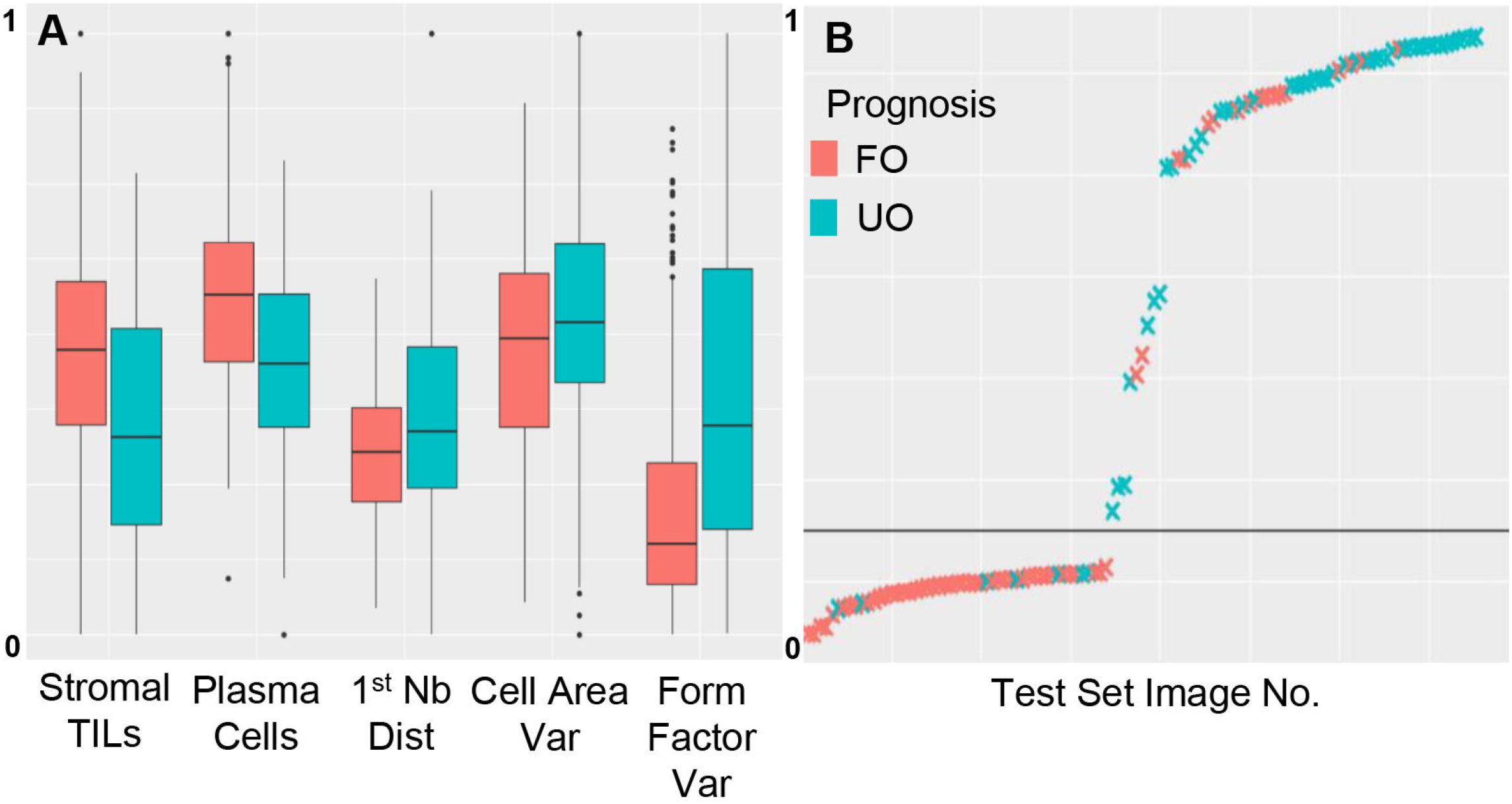
(A) Boxplots of selected prognostic variables identified with the workflow. Values have been normalised within a range of 0 to 1. (B) The probability of each image being from an UO patient as predicted by the neural network model, arranged from lowest to highest probability when applied to the test set.

We trained a neural network (NN) to use all statistically significant variables to predict patient outcomes. Fig 3B shows the NN applied to the test set of images and a clear split between the UO and FO groups. The model had an overall accuracy of 78.76%. In predicting patients with FO, it had a sensitivity of 72.13% and a specificity of 86.54%. The model had a Kappa statistic of 0.579, indicating a good model considering the difficulty of the task^22^.

Furthermore, a k-fold cross-validation was used to validate the model using 10 folds. This revealed similar results when the model was applied to the test set with an average overall accuracy of 77.72% and Kappa statistic of 0.552. The average sensitivity and specificity for the favourable outcome group was 77.17% and 78.12% respectively.

## Discussion

Recent years have seen a growing number of calls to de-intensify treatment regimens for HPV+OpSCC since the majority of these patients demonstrate better overall and disease-specific survival compared to HPV-unrelated counterparts. However, a significant proportion of patients with HPV+OpSCC still exhibit UO following radical treatment for whom de-intensification would be inappropriate^5^. While various biomarkers have proposed to have prognostic value in HPV+OpSCC, none of these have been validated for routine clinical practice or are under investigation to stratify patients in de-escalation trials. Against this background, routine diagnostic H&E sections contain a wealth of data which can be exploited for prognostic purposes without the need for additional laboratory tests which may be costly and delay the initiation of treatment. However, assessment of H&E features by pathologists is challenging, subject to bias and lacks reproducibility. For example, in Fig 1C, representative photomicrographs from patients with FO (C1) and UO (C2) show similar histological features, illustrating the challenges of subjective prognostication, even to an experienced pathologist. By contrast, HCA and various implementations of machine learning algorithms have been successfully employed to evaluate histology images in cancers^23^, but these tools are currently unavailable for OpSCC. Therefore, in the present work we aimed at building an automated tool to analyse large numbers of histological images accurately and reproducibly, to demonstrate the value of our tool to detect and measure prognostic features, and to predict patient outcomes in HPV+OpSCC.

Our work has identified various prognostic factors, some of which corroborates with established findings in the literature while others are fairly novel. TILs have been shown to be a prognostic factor in HPV+OpSCCs^10^. However, many studies employ a semi-quantitative method of assessment with suboptimal inter-observer agreement and reproducibility. A digital workflow is able to perform the exact same steps with each round of analysis. Furthermore, such methods of assessment are time-consuming and labour-intensive for the quantification of a single prognostic feature. Many patients exhibit a strong immune infiltrate consisting of various immune cell types like plasma cells and TILs and this complicates the current proposed methods of TIL quantification by estimating the area of stroma covered by TILs^24^. Furthermore, to our best knowledge, previous studies have not investigated the prognostic value of TILs in different tissue compartments, stromal versus intratumoural in HPV+OpSCC. Our various measurements of TILs have all shown to be strong prognostic factors. The implementation of the TIL index to account for variation in area of tumour to stroma resulted in a high statistical significance, improving our confidence in the measurement of this variable as a prognostic feature.

We found that stromal plasma cells were increased in patients with FO. Few studies have investigated the role of plasma cells in the stromal component of HPV+OpSCCs and many studies choose to ignore this subset of immune cells. However, some recent work has suggested plasma cells may exhibit resistance to radiotherapy over other B cells^25^. This may have an impact on the antigen-specific antibody response to the tumour but further studies elucidating the biological role of plasma cells in HPV+OpSCCs are required. Another study looking at a plasma cell marker, immunoglobulin J polypeptide (IGJ), found that patients with higher IGJ had better disease-specific and overall survival^12^. While none of these studies directly quantify plasma cells in H&E slides, they are in agreement with the trends we have observed, and we show an alternative method to quantify plasma cells as a prognostic feature without the transcriptomics or flow cytometry used in these studies.

Our workflow has managed to automate the single-cell quantification of anisonucleosis and nuclear pleomorphism from several morphological measurements of tumour nuclei.Previously, Lewis and colleagues reported an association between cellular anaplasia and UO in HPV+OpSCCs^26^. Similarly, our results revealed patients with FO tended to have rounder, less eccentric nuclei with smaller perimeters for the same area. Interestingly, while the size of the nuclei was not significantly different between the two groups when measured across thousands of nuclei, there was greater heterogeneity in the sizes of nuclei from UO patients. This highlights the importance of intratumour heterogeneity as prognostic feature, while the average of a measured variable may not be prognostic itself, the degree of spread of the data within the quantified variable may be prognostic. UO patients had greater intratumour heterogeneity in all 5 morphological measurements with 4 of them being statistically significant by univariate analysis. To the best of our knowledge, quantification of morphological intratumour heterogeneity has not been reported in the literature for HPV+OpSCCs. This morphologic heterogeneity may represent an underlying genetic heterogeneity in patients with UO, although further investigations are required to identify this potential link.

A previous study by Ali and colleagues found that the spatial distribution and tumour nuclear clustering was helpful in developing a predictive model of outcome in p16+ OpSCC patients^27^. Similarly, we found that tumour nuclei from FO patients clustered closer to one another as evidenced by higher number of neighbours and shorter first and second neighbour distances. Furthermore, this pattern of spatial distribution was more homogenous across the tumour for these FO patients. In non-HPV head and neck cancer, Delides and colleagues quantified the heterogeneity of spatial distribution of laryngeal carcinoma by measuring fractal dimensions^28^. Their work revealed that patients with poor outcomes had neoplastic tissues that exhibited greater spatial irregularities.

Texture features quantified by image analysis have been used to prognosticate patients from various cancer types. However, most studies measure textural features across the whole image rather than within the individual nuclei, making it more difficult to elucidate their biological significance. However, a study in breast cancer measured nuclear textural features as a way to quantify DNA arrangement and found some texture measurements to improve the prognostication of patients^29^. Our study similarly found several texture and granularity measurements within tumour nuclei to be prognostic and these features were used to train our neural network model. Furthermore, we attempted to quantify distinct nucleoli by searching for small, round objects within tumour nuclei. We found patients with more nucleoli had better outcomes and as expected, these patients also had higher textural and granularity values. This shows that while textural features can be difficult to interpret and ascribe biological significance to, they may be used to measure visual and biologically relevant features within tumour cells. The presence and size of nucleoli have been reported to carry both favourable and unfavourable prognosis in different cancer types^30,31^. Hence, we interpret these findings with caution as it is currently not possible to determine the quality of function of these nucleoli through image analysis alone.

Overall, our model captures all these features providing ground to build predictive statistical models able to account for multiple prognostic features. Importantly, most of our selected features have direct link with known biological counterparts, thus our HCA tools could be used to develop new mechanistic hypotheses and computational models to complement and enrich statistical inference. Nonetheless, none of these prognostic features are able to predict patient outcomes on their own. There is significant inter-sample heterogeneity as visualised in the heat map (Fig 2.) and despite the patients generally clustering by outcome, they do not form groups that are easily separable. Hence, AI tools that take into account multiple variables and large quantities of data are required to make accurate predictions of patient prognoses. In this study, we have shown the usefulness of such tools and their ability to achieve promising accuracies.

We show that the development of a predictive statistical model is possible by automated quantification of routine H&E biopsies without staining for additional biomarkers or other diagnostics. This is also done in a timely manner, avoiding delays to treatment initiation. The entire workflow averaged under an hour of processing time per patient on a standard computer, without the need to purchase dedicated computational resources. While we have developed a workflow that does not require the additional time and cost involved in obtaining whole-slide scans, our platform is also easily adaptable to analyse whole-slide scans which might be advantageous in a diagnostic digital pathology service context. Such a workflow using free-to-use, open-source software is easy to implement, without high financial barriers to entry. This allows a digital pathology workflow to be implemented even in developing countries to improve diagnostic capabilities.

## Conclusion

In summary, we built a single-cell quantitative image analysis workflow to analyse HPV+OpSCC H&E slides to identify prognostic factors and quantify their heterogeneity within the tumour. We have shown that some of these measures are predictive in their own right, and that their variance within a tumour can itself be prognostic and contribute strongly to the accuracy of predictive models.

Our open-source high content analysis workflow on routine H&E slides and statistical modelling can aid prognostication of HPV+OpSCCs with promising accuracy. Our work supports the use of machine learning-powered high content analysis followed by statistical modelling in digital pathology to exploit clinically relevant features in routine diagnostic pathology without additional laboratory diagnostics.

## Data Availability

All data produced in the present study are available upon reasonable request to the authors.

https://github.com/jonashue1/HPV-OpSCC-Machine-Learning

## Acknowledgements

This research did not receive any specific grant from funding agencies in the public, commercial, or not-for-profit sectors.

